# Missed opportunities for hospitalisation of severe pneumonia identified using Integrated Management of Childhood Illness and pulse oximetry at primary care among children aged 2 to 59 months: the AIRE research cohort, 2021 - 2022

**DOI:** 10.1101/2025.02.03.25321599

**Authors:** Dahourou Désiré Lucien, Néboua Désiré, Hedible Gildas Boris, Zair Zineb, Louart Sarah, Guilavogui Jean Paul Yassa, Sawadogo Abdoul Guaniyi, Meda Bertrand, Ouédraogo Yugbaré Solange, Diallo Ibrahima Sory, Diakité Abdoul Aziz, Abarry Souleymane Hannatou, Ridde Valéry, Leroy Valériane, the AIRE Research Study Group

**Affiliations:** Département Biomédical et de Santé Publique, Institut de Recherche en Sciences de la Santé (IRSS/CNRST), Ouagadougou, Burkina Faso; Center for Epidemiology and Research in Population Health (CERPOP), UMR 1295, Inserm, University Paul Sabatier Toulouse 3, Toulouse, France; ALIMA, The Alliance for International Medical Action, Dakar, Senegal; University of Lille, CLERSE – Centre Lillois d’Études et de Recherches Sociologiques et Économiques, Lille, France; ALIMA, The Alliance for International Medical Action, Conakry, Guinea; Tdh, Terre des hommes, Ouagadougou, Burkina Faso; Solthis, Niamey, Niger; CHU de Bogodogo, Ouagadougou, Burkina Faso; Institut de Nutrition et Santé de l’Enfant (INSE), Conakry, Guinée; CHU Gabriel Touré, Bamako, Mali; Ministère de la santé, des populations et des affaires sociales, Niamey, Niger; Université Paris Cité, IRD, Inserm, Ceped, F-75006 Paris, France

**Author notes:** See composition in the appendix section. Corresponding author: Valériane Leroy, Center for Epidemiology and Research in Population Health (CERPOP), UMR 1295, Inserm, University Paul Sabatier Toulouse 3, Toulouse, France.

**Keywords:** IMCI guidelines, severe pneumonia, hypoxemia, hospital referral, child health

## Abstract

**Background:** The Integrated Management of Childhood Illness (IMCI) guidelines guide the healthcare workers (HCW) referral decision of under-5 children at primary health centres (PHCs). The AIRE project has implemented the routine Pulse Oximeter (PO) use into IMCI consultations at PHCs in West Africa (Burkina Faso, Guinea, Mali and Niger) to improve the diagnosis and referral of severe cases. We analysed the frequency and correlates of missed opportunities for hospitalisation (MOH) of IMCI severe pneumonia cases.

**Methods:** All the children aged 2-59 months attending IMCI consultations, classified as severe cases using IMCI+PO were enrolled with parental consent in a prospective cohort of 14-day in 16 research PHCs (4/country). HCW referral decision, and MOH for severe pneumonia were described. Correlates of MOH were investigated using a generalized linear mixed regression model with a random effect on country.

**Results:** From June 2021 to June 2022, among the 1,786 children aged 2-59 months classified as severe cases by IMCI+PO, 682 (38.2%) were severe pneumonia: among those, 35 (5.1%) had also a severe anaemia, 47 (6.9%) a severe hypoxemia (SpO_2_<90%) and 602 (88.3%) a severe malaria. Overall, HCW made the referral decision for 125 (18.3%) children, refused by three (2.4%) families; thus, 560 (82.1%) were MOH. Severe anaemia reduced the adjusted odds ratio [aOR] of MOH (aOR: 0.02; 95% confidence interval [95%CI]: 0.01-0.07) while having an SpO_2_ between 90 to 93% (aOR: 12.16, 95%CI: 3.47-42.61), or greater than 94% (aOR: 11.81; 95%CI: 3.98-35.02), or a severe malaria (aOR: 2.55; 95%CI: 1.04-6.26) significantly increased it.

**Conclusion:** MOH for severe pneumonia were extremely high at PHC level in these settings, mainly explained by HCW’ decisions. The absence of severe hypoxemia was an expected decision factor but co-morbidity with severe malaria is a concern. Supporting HCW and hospital referral remain essential to improve the management of severe pneumonia.

*What is already known on this topic:*

- Pneumonia remains the leading cause of under-5 children mortality in sub-Saharan Africa.
- The implementation of standardized guidelines, such as the World Health Organization’s (WHO’s) Integrated Management of Childhood Illnesses (IMCI) has reduced mortality from pneumonia but remain insufficient.
- Introduction of Pulse Oximetry at primary care should improve the diagnosis of severe hypoxemia associated with severe pneumonia requiring urgent referral.
- Little is known about the referral to hospital and hospitalisations of severe pneumonia cases identified in primary care in West African settings.

*What this study adds:*

- This study, carried out in 16 primary health care centres in four West African countries (Burkina Faso, Guinea, Mali, Niger) showed a high rate of severe pneumonia among severe IMCI cases (38%) and a high rate of missed opportunities for hospitalisation of these severe pneumonia (82.1%). This latter was mainly attributed to the lack of referral decision made by healthcare workers (HCW).
- It is notifiable that co-morbidity with severe malaria was also frequent, 88%.
- The absence of severe hypoxemia (SpO_2_<90%) and of severe anaemia reduced the risk of missed opportunity of hospital referral but severe malaria co-morbidity increased it.

*How this study might affect research, practice or policy:*

- The use of PO was associated with a more appropriate HCW’ referral decision for severe pneumonia with severe hypoxemia, but the management of co-morbidity with severe malaria should be strengthen to improve optimal care.
- Decision makers should be sensitized on the importance of missed opportunities for hospital referral of severe pneumonia, which remains the leading killer of under-5 children, and mobilised to improve their hospital referral system with appropriate management to improve under-5 children quality of care.

## Introduction

Globally, there are over 1,400 cases of pneumonia per 100,000 children, with the greatest incidence occurring in South Asia (2,500 cases per 100,000 children) and West and Central Africa (1,620 cases per 100,000 children) [1]. Pneumonia is one of the leading causes of death among under-5 children. In 2019, there were 45 million episodes of pneumonia in under-5 children worldwide, leading to more than 700,000 deaths [2]. Several risk factors for pneumonia incidence and severity have been identified, including child age, lack of immunisation, malnutrition, chronic underlying diseases, HIV infection or HIV exposure in young infants, young maternal age, low maternal education, low socio-economic status, and smoke exposure or indoor air pollution [3].

An effective management is crucial for reducing pneumonia-related morbidity and mortality in children [4]. The World Health Organization (WHO) has developed Integrated Management of Childhood Illnesses (IMCI) guidelines at primary care and many low– and middle-income countries (LMICs) used this syndromic approach to guide healthcare workers (HCWs) decision in disease classification and management of under-5 children [5]. More specifically, the IMCI proposes a comprehensive approach to the management of childhood illnesses including pneumonia, with prompt assessment and classification, appropriate treatment with antibiotics, supportive care and hospital referral for severe cases. In 2012, the WHO revised its pneumonia management guidelines. Children aged 2–59 months presenting with a cough or breathing difficulties with fast breathing or chest indrawing are classified as “pneumonia” and should receive oral amoxicillin with home care advice. While children aged 2–59 months with cough or difficult breathing with at least one general danger signs (not able to drink, persistent vomiting, convulsions, lethargic or unconscious, stridor in a calm child or severe malnutrition) are classified as “severe pneumonia or very severe disease”. In this case, the recommended management is to administer a first dose of antibiotic and urgently refer the patient to hospital for injectable antibiotics and supportive therapy, including oxygen therapy if severe hypoxemia is diagnosed using a Pulse Oximeter (PO) [6]. When applied with high fidelity, IMCI-guidelines may improve health outcomes [7]. However, data about hospitalisation rates and associated factors, including HCWs’ adherence to referral guidelines for severe IMCI-related pneumonia are lacking at primary care level.

From 2021 to 2022, the AIRE (Améliorer l’Identification des détresses Respiratoires chez l’Enfant) operational project has implemented the routine use of PO in IMCI consultations for under-5 children to improve the diagnosis of severe hypoxemia and its management at primary health care centres (PHCs) in Burkina Faso, Guinea, Mali and Niger [8]. Several papers are reported elsewhere on the context, process, and children’s outcomes after implementation of PO use in this project [9–12]. In this paper, we specifically aimed to describe the frequency, associated factors and outcomes of the missed opportunities for hospitalisation of cases of IMCI-related severe pneumonia identified in the research PHCs of the four AIRE countries.

## Methods

### Study sites

This analysis used individual data from the AIRE project conducted by a consortium of three NGOs (ALIMA, Solthis, Terre des hommes) and the French Institute of Health and Medical Research (Inserm). Briefly, the AIRE project has introduced the routine PO use into IMCI consultations in 202 PHCs in the above four West African countries [8]. At baseline, all HCWs were trained or recycled in national IMCI protocols integrating PO. From June 14^th^, 2021 to June 19^th^, 2022, a research operational study was conducted in the 16 selected research public PHCs (4 per country) and 8 district referral hospitals in the four countries to evaluate the implementation of PO use into IMCI guidelines [11,12,13].

### Study design and inclusion criteria and procedure

All outpatient children aged 0-59 months attending IMCI consultations at the 16 research PHCs were eligible for PO use except children aged 2-59 months classified as simple cases and without cough or breathing difficulties. During the consultation, the PHC-based HCW classified children into three groups using the IMCI classification: green for simple cases, yellow for moderate cases and red for severe cases, then used PO. All the children aged 0-59 months classified as severe cases were consecutively enrolled over 12 months in a prospective cohort and followed until Day-14, with written parental consent. Children were not included on nights, weekends and public holidays because the research teams were not always on site. Children aged 2-59 months and classified as severe pneumonia were enrolled in this specific analysis.

### Data collection and definitions

At all research sites, data were collected using an electronic case report form developed with REDCap software by dedicated data collectors at PHC and hospital levels. Sociodemographic and clinical individual data, including IMCI classifications, clinician decision for hospital referral at the PHC level and actual hospitalisation were collected.

The WHO’s revised pneumonia management guidelines define severe pneumonia in children aged 2–59 months with cough or difficult breathing, and with any general danger signs (not able to drink, persistent vomiting, convulsions, lethargic or unconscious) or stridor in a calm child [6]. Fast breathing is defined as a respiratory rate greater than or equal to 50 breaths per minute for children aged from 2 to 11 months inclusive and greater than or equal to 40 breaths per minute for those aged 12 to 59 months inclusive [14]. Normal heart rate is defined as heart rate between 100 and 160 per minute in children aged from 0 to 1 year, from 90 to 150 for children from 1 to 3 years, and from 80 to 140 for those aged between 3 to 5 years [15]. In this study, severe hypoxemia was defined as an SpO_2_ level less than 90% and moderate hypoxemia as an SpO_2_ level between 90% and 93% [16,17].

### Outcomes

The WHO revised pneumonia management guidelines recommended the urgent referral to hospital for severe IMCI-pneumonia, right after a first dose of antibiotic given at PHC [6]. The existing IMCI guidelines were all in line with this recommendation in the four AIRE countries. The study outcome was the missed opportunity for hospitalisation of severe pneumonia cases diagnosed at PHC. This is defined as all severe pneumonia cases who should be referred [6], but were not referred for any reason after the IMCI consultation.

### Statistical analysis

Proportions of children with severe pneumonia, their socio-demographics and clinical characteristics, and their vital status at day-14 were described and compared according to their actual hospitalisation. Quantitative data were described using means and standard deviations or using median and interquartile ranges and compared using Student’s t-tests. Categorial data were described with proportions and were compared using Pearson χ2 or Fischer exact tests. All analysis were considered statistically significant with p-value less than 0.05.

We analysed the factors associated with the missed opportunities for hospitalisation, using a generalized linear mixed regression model with a random effect on country. We computed a global analysis including all participants, and a specific analysis for participants enrolled in Guinea where the majority of severe pneumonia cases were enrolled. For each analysis, we computed an explanatory model with univariate analysis, then adjusted a full model including all the relevant variables or those associated with a p-value <0.20 in univariate analysis. We report adjusted Odds Ratio (aOR) with their 95% confidence Intervals (95%CI). Variables explored were: mother’s vital status and education level, carrying on an income-generating activity, travel time from home to PHC (>30 min), consultation delay (>2 days) since the onset of disease, type of IMCI support (paper or electronic), SpO_2_ level, presence of co-morbidity (severe malaria, severe anaemia and severe malnutrition). Age and sex were forced in multivariate models. A two-tailed p-value of <0.05 was regarded as statistically significant. R software version 4.0.5 was used for all analysis.

### Ethical aspects

The AIRE research protocol, has been approved by each national ethics committee, by the Inserm Institutional Evaluation Ethics Committee (IEEC) and the WHO Ethics Review Committee (WHO-ERC): Comité d’Ethique pour la Recherche en Santé (CERS), Burkina Faso n°2020-4-070; Comité National d’Ethique pour la Recherche en Santé (CNERS), Guinea n°169/CNERS/21; Comité National d’Éthique pour la Santé et les Sciences de la vie (CNESS), Mali n°127/MSDS-CNESS; Comité National d’Ethique pour la Recherche en Santé (CNERS) Niger n°67/2020/CNERS; Inserm IEEC n°20-720; WHO-ERC n° ERC.0003364). This study has been retrospectively registered in the Pan African Clinical Trials Registry on June 15th 2022 under the following Trial registration number: PACTR202206525204526 [8].

### Patient and public involvement

This study was conducted using individual data collected with the ethical committees’ and Ministries of Health’ (MOHs) authorization. Patients were not involved in the design, or conduct, or reporting, or dissemination plans of our research.

## Results

From June 14^th^, 2021 to June 20^th^, 2022, 39,360 under-5 children attended IMCI consultations in the 16 research PHCs. Among them, the 7,760 (19.7%) simple non-respiratory IMCI cases not eligible for PO use were excluded. Among the 31,600 (80.3%) children eligible for PO use, 15,670 (49.6%) seeking health services at night or over the weekends were not invited to participate into the study. Among the 15,930 (50.4%) remaining who were invited to participate, 33 (0.2%) families refused, mainly because the child’s accompanying person needed the father’ consent, and 15,897 (99.7%) were included in the research study with parental consent, of whom 61 (0.4%) were excluded from the analysis for missing IMCI classification or wrong inclusion. A total of 15,836 under-5 children were included in the study, of whom 1,998 were classified as severe cases using IMCI+PO. Those included 212 (10.6%) neonates aged <2 months, excluded from this analysis (Figure 1). Of the 1,786 children aged 2-59 months, 682 children were classified as severe pneumonia, and were enrolled in this analysis. The prevalence of severe pneumonia was 38.2% (682/1,786) with large heterogeneity between countries: 5.6% (38/196) in Burkina Faso, 73.8% (501/600) in Guinea, 12.8% (69/707) in Mali and 26.1% (74/283) in Niger.

**Figure 1.**
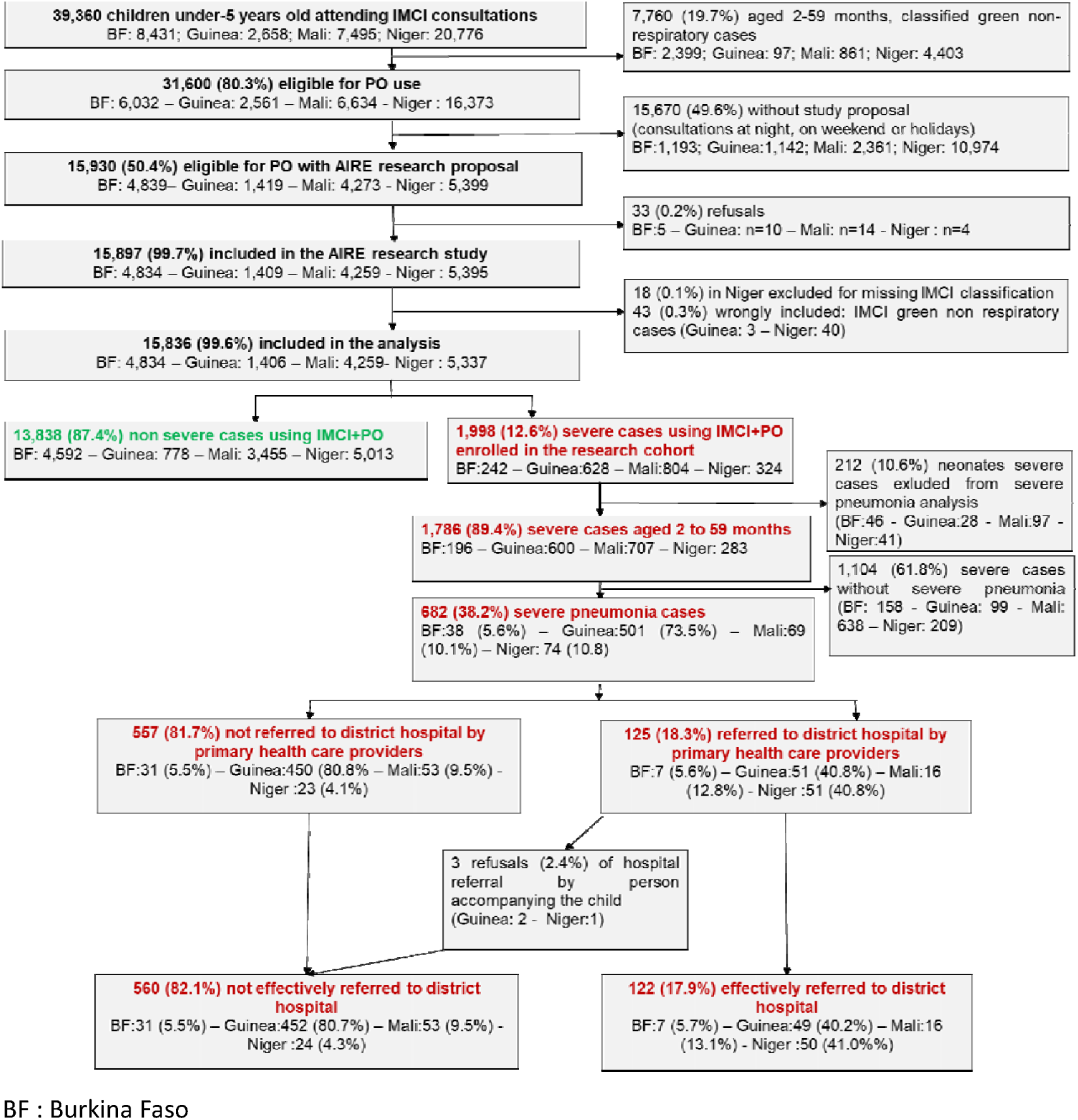
Flowchart of the inclusion process of children 2-59 months of age enrolled in the pneumonia sub-study of missed opportunity of hospital referral in the AIRE research study, June 2021 – June 2022.

### Baseline characteristics and missed opportunities of hospitalisation

Among the 682 severe pneumonia cases enrolled, median age was 23.2 months (interquartile range: 12.1-36.4 months); 35 (5.1%) had also a severe anaemia, 47 (6.9%) a severe hypoxemia (SpO_2_<90%) and 602 (88.3%) a severe malaria. Overall, HCW took the referral decision for 125 (18.3%) children, but three (2.4%) families refused it, and 557 (81.7%) were not referred to district hospital by HCW. Overall, 560 (82.1%) were missed opportunities for hospitalisation: 31 (5.5%) from Burkina Faso, 452 (80.7%) from Guinea, 53 (9.5%) from Mali, and 24 (4.3%) from Niger (Figure 1).

Table 1 presents the characteristics of the 682 severe pneumonia cases enrolled according to their actual hospitalisation. Cases not hospitalised were significantly older than those hospitalised (p<0.001). There were differences in hospitalisation rates of severe pneumonia according to head of household education level, income generating activity of the person accompanying the child, travel time from home to PHC, clinical symptoms, hypoxemia, severe malaria, severe anaemia and severe malnutrition. The proportion of the head of household who never attended school (59.8%, 73/122) was significantly higher for children hospitalised compared to those not hospitalised (52.3%, 293/560). The proportion of the child accompanying person who did not have an income-generating activity (73.8%, 90/122) was significantly higher for children hospitalised compared to those not hospitalised (55,9%, 313/560). Compared to children not hospitalised (72.1%, 404/560), those hospitalised had a significantly shorter travel time from home to PHC (58.2%, 71/122). The proportions of children with tachycardia (38.5%, 47/122), fast breathing (40.2%, 49/122) and chest indrawing (18.9%, 23/122) were significantly higher among those hospitalised compared to those not hospitalised. The proportions of children with hypoxemia (32%, 39/122), severe anaemia (22.1%, 27/122) and who were severely malnourished (15.3, 15/122) were significantly higher among those hospitalised compared to those not hospitalised. However, the proportion of severe malaria (63.1%, 77/122) was significantly lower among those hospitalised compared with those not hospitalised (93.8%, 525/560).

**Table 1.** Baseline characteristics of the severe pneumonia cases aged 2-59 months enrolled in the AIRE project according to their hospitalisation, June 2021 – June 2022. (N=682)

| Variables |  | Hospitalisation after IMCI consultation |  | p value | Total |
| --- | --- | --- | --- | --- | --- |
|  |  | Yes | No |  |  |
|  |  | N=122 | N=560 |  | N=682 |
| **Country** | Burkina Faso, n (%) | 7 (5.7) | 31 (5.5) | <0.001 | 38 (5.6) |
|  | Guinea, n (%) | 49 (40.2) | 452 (80.7) |  | 501 (73.5) |
|  | Mali, n (%) | 16 (13.1) | 53 (9.5) |  | 69 (10.1) |
|  | Niger, n (%) | 50 (41.0) | 24 (4.3) |  | 74 (10.8) |
| **Age groups, in months** | [2 to 23], n (%) | 86 (70.5) | 285 (50.9) | <0.001 | 371 (54.0) |
|  | [24 to 59], n (%) | 36 (29.5) | 275 (49.1) |  | 311 (46.0) |
| **Sex** | Female, n (%) | 50 (41.0) | 281 (50.2) | 0.082 | 331 (49.0) |
| **Dead mother** | n (%) | 1 (0.8) | 0 (0.0) | 0.402 | 1 (0.2) |
|  | Missing, n (%) | 0 (0.0) | 0 (0.0) |  | 0 (0.0) |
| **Education level of head of household** | Never attended school, n (%) | 73 (59.8) | 293 (52.3) | 0.0321 | 366 (54.0) |
|  | Missing, n (%) | 0 (0.0) | 0 (0.0) |  | 0 (0.0) |
| **Person accompanying the child** | Mother, n (%) | 118 (96.7) | 542 (96.8) |  | 660 (97.0) |
|  | Father, n (%) | 1 (0.8) | 4 (0.7) | 0.992 | 5 (0.7) |
|  | Others, n (%) | 3 (2.5) | 14 (2.5) |  | 17 (2.5) |
|  | Missing, n (%) | 0 (0.0) | 0 (0.0) |  | 0 (0.0) |
| **Ability to read or write of the person accompanying the child** | No, n (%) | 89 (73.0) | 358 (63.9) | 0.073 | 447 (65.5) |
|  | Missing, n (%) | 0 (0.0) | 0 (0.0) |  | 0 (0.0) |
| **Income-generating activity of person accompanying the child** | No, n (%) | 90 (73.8) | 313 (55.9) | <0.001 | 403 (59.1) |
|  | Missing, n (%) | 0 (0.0) | 0 (0.0) |  | 0 (0.0) |
| **Electronic IMCI support** | Yes, n (%) | 15 (12.3) | 74 (13.2) | 0.900 | 89 (13.0) |
|  | Missing, n (%) | 0 (0.0) | 0 (0.0) |  | 0 (0.0) |
| **Travel time Home - PHC** | > 30 min, n (%) | 51 (41.8) | 156 (27.9) | 0.003 | 207 (30.4) |
|  | Missing, n (%) | 0 (0.0) | 0 (0.0) |  | 0 (0.0) |
| **Consultation delay since onset of signs in days** | > 2 days, n (%) | 84 (68.9) | 340 (60.7) | 0.239 | 424 (62.2) |
|  | Missing, n (%) | 10 (8.2) | 16 (2.9) |  | 26 (3.8) |
| **Temperature** | (T> 38°C), n (%) | 23 (18.9) | 92 (16.4) | 0.189 | 115 (16.9) |
|  | Missing, n (%) | 68 (55.7) | 275 (49.1) |  | 343 (50.3) |
| **Heart rate for age** | Bradycardia, n (%) | 8 (6.6) | 7 (1.3) | <0.001 | 15 (2.2%) |
|  | Normal, n (%) | 52 (42.6%) | 348 (62.1) |  | 400 (58,6) |
|  | Tachycardia, n (%) | 47 (38.5) | 199 (35.5) |  | 246 (36,1) |
|  | Missing, n (%) | 15 (12.3) | 6 (1.1) |  | 21 (3.1) |
| **Respiratory rate for age** | Slow breathing, n (%) | 1 (0.8) | 0 (0.0) | <0.001 | 1 (0.1) |
|  | Normal breathing, n (%) | 41 (33.6) | 351 (62.7) |  | 392 (57,5) |
|  | Fast breathing, n (%) | 49 (40.2) | 139 (24.8) |  | 188 (27,6) |
|  | Missing, n (%) | 31 (25.4) | 70 (12.5) |  | 101 (14.8) |
| **Chest indrawing** | No, n (%) | 99 (81.1) | 534 (95.4) | <0.001 | 633 (92.8) |
|  | Yes, n (%) | 23 (18.9) | 26 (4.6) |  | 49 (7.2) |
|  | Missing, n (%) | 0 (0.0%) | 0 (0.0%) |  | 0 (0.0%) |
| **SpO<sub>2</sub> level** | Severe hypoxemia SpO<sub>2</sub><90%, n (%) | 39 (32.0) | 8 (1.4) | <0.001 | 47 (6.9) |
|  | Moderate hypoxemia SpO<sub>2</sub> [90-93%], n (%) | 13 (10.7) | 54 (9.6) |  | 67 (9.8) |
|  | No hypoxemia SpO<sub>2</sub>≥94%, n (%) | 69 (56.6) | 496 (88.6) |  | 565 (82.8) |
|  | Missing, n (%) | 1 (0.8) | 2 (0.4) |  | 3 (0.4) |
| **Severe malaria** | Yes, n (%) | 77 (63.1) | 525 (93.8) | <0.001 | 602 (88.3) |
| **Severe anaemia** | Yes, n (%) | 27 (22.1) | 8 (1.4) | <0.001 | 35 (5.1) |
| **Severe malnutrition** | Yes, n (%) | 15 (12.3) | 22 (3.9) | <0.001 | 37 (5.4) |

### Factors associated with missed opportunities for hospitalisation of severe pneumonia

Table 2 presents a multivariate generalized linear mixed regression model of associated factors with missed opportunities for hospitalisation for all severe pneumonia. Adjusted for age and sex, income generating activity of the person accompanying the child, travel time from home to PHC, and type of IMCI used, SpO_2_ level, severe malaria and severe anaemia were independently associated with missed opportunity of hospitalisation. The odds of not being hospitalised were significantly higher for children with moderate hypoxemia (SpO_2_ between 90 to 93%, adjusted odds ratio [aOR]: 12.16, 95% confidence interval [95%CI]: 3.47-42.61), and those with no hypoxemia (SpO_2_ greater than 94%, (aOR: 11.81; 95%CI: 3.98-35.02) compared with children with severe hypoxemia (SpO_2_<90%). The odds of not being hospitalised were significantly lower for children with severe anaemia (aOR: 0.03; 95%CI: 0.01-0.07) compared to those without severe anaemia. However, the odds of not being hospitalised were significantly higher for children with severe malaria (aOR: 2.55; 95%CI: 1.04-6.26) compared to those without severe malaria.

**Table 2.** Factors associated with the missed opportunities for hospitalisation of severe pneumonia cases in children aged 2-59 months in all countries, generalized linear mixed regression model (N=682)

| Variables |  | Missed opportunities for hospitalisation |  | Univariate | Adjusted analysis |
| --- | --- | --- | --- | --- | --- |
|  |  | Yes | No | OR (95% CI, p value) | aOR (95% CI, p value) |
| Age (months) | [2 - 23] | 285 (50.9) | 86 (70.5) | Reference |  |
|  | [24 - 59] | 275 (49.1) | 36 (29.5) | 1.66(1.02-2.68, p=0.04) | 1.52(0.83-2.76, p=0.172) |
| Sex | Female | 281 (50.2) | 50 (41.0) | Reference |  |
|  | Male | 279 (49.8) | 72 (59.0) | 0.69(0.44-1.08, p=0.104) | 0.78(0.45-1.35, p=0.373) |
| Ability to read or write of the person accompanying the child | No | 358 (63.9) | 89 (73.0) | Reference |  |
|  | Yes | 202 (36.1) | 33 (27.0) | 1.54(0.85-2.78, p=0.157) | 1.24(0.6-2.57, p=0.557) |
| Income-generating activity of person accompanying the child | No | 313 (55.9) | 90 (73.8) | Reference |  |
|  | Yes | 247 (44.1) | 32 (26.2) | 1.22(0.72-2.06, p=0.465) |  |
| Travel time Home - PHC | >30mn | 156 (27.9) | 51 (41.8) | Reference |  |
|  | ≤30mn | 404 (72.1) | 71 (58.2) | 2.56(1.49-4.41, p=0.001) | 1.55(0.81-2.96, p=0.185) |
| Type of IMCI Support | Papier-based | 486 (86.8) | 107 (87.7) | Reference |  |
|  | Electronic-based | 74 (13.2) | 15 (12.3) | 4.82(0.87-26.86, p=0.072) | 2.05(0.45-9.34, p=0.353) |
| Consultation delay since the onset of symptoms (days) | >2 days | 324 (59.6) | 74 (66.1) | Reference |  |
|  | ≤2 days | 220 (40.4) | 38 (33.9) | 1.67(1.01-2.75, p=0.044) | 1.21(0.68-2.18, p=0.515) |
|  | Yes | 411 (73.4) | 106 (86.9) | 0.72(0.37-1.41, p=0.341) |  |
| SpO<sub>2</sub> level | <90% | 8 (1.4) | 39 (32.2) | Reference |  |
|  | [90,93%] | 54 (9.7) | 13 (10.7) | 19.27(5.94-62.49, p<0.001) | 12.16(3.47-42.61, p<0.001) |
|  | ≥94% | 496 (88.9) | 69 (57.0) | 22.82(8.41-61.91, p<0.001) | 11.81(3.98-35.02, p<0.001) |
| Severe malaria | No | 35 (6.2) | 45 (36.9) | reference |  |
|  | Yes | 525 (93.8) | 77 (63.1) | 2.39(1.12-5.1, p=0.024) | 2.55(1.04-6.26, p=0.041) |
| **Severe anaemia** | No | 552 (98.6) | 95 (77.9) | reference |  |
|  | Yes | 8 (1.4) | 27 (22.1) | 0.02(0.01-0.06, p<0.001) | 0.03(0.01-0.07, p<0.001) |
| **Severe acute malnutrition** | No | 517 (95.9) | 103 (87.3) | reference |  |
|  | Yes | 22 (4.1) | 15 (12.7) | 1.14(0.44-2.93, p=0.783) |  |

OR : odds ratio; AOR : adjusted odds ratio

We also carried out a sensitivity analysis of the model restricted to severe pneumonia in Guinea, accounting for 80.7% of all cases. In Guinea, adjusted for age, sex, travel time from home to PHC, level of SpO_2_ and severe malnutrition, only severe anaemia was independently associated with missed opportunities of hospitalisation. The odds of not being hospitalised were significantly lower for children with severe anaemia (aOR: 0.02; 95%CI: 0.01-0.07) compared to those without severe anaemia (table 3).

**Table 3.** Factors associated to missed opportunities for hospitalisation of severe pneumonia cases in Guinea, generalized linear mixed regression model. (N=501)

| Variables |  | Missed opportunities for hospitalisation |  | Univariate | Adjusted analysis |
| --- | --- | --- | --- | --- | --- |
|  |  | Yes | No | OR (95% CI, p value) | aOR (95% CI, p value) |
| Age (months) | [2 - 23] | 217 (48.0) | 32 (65.3) |  |  |
|  | [24 - 59] | 235 (52.0) | 17 (34.7) | 1.96(1.05-3.65, p=0.034) | 1.67(0.78-3.6, p=0.187) |
| Sex | Female | 223 (49.3) | 25 (51.0) |  |  |
|  | Male | 229 (50.7) | 24 (49.0) | 1.03(0.57-1.86, p=0.929) | 0.92(0.44-1.91, p=0.818) |
| Dead mother | Yes | 0 (0.0) | 0 (0.0) |  |  |
|  | No | 452 (100.0) | 49 (100.0) |  |  |
| Ability to read or write of the person accompanying the child | No | 265 (58.6) | 28 (57.1) |  |  |
|  | Yes | 187 (41.4) | 21 (42.9) | 1.53(0.73-3.17, p=0.258) |  |
| Income-generating activity of person accompanying the child | No | 225 (49.8) | 28 (57.1) |  |  |
|  | Yes | 227 (50.2) | 21 (42.9) | 1.28(0.7-2.36, p=0.422) |  |
| Travel time Home - PHC | >30mn | 125 (27.7) | 21 (42.9) |  |  |
|  | ≤30mn | 327 (72.3) | 28 (57.1) | 2.18(1.17-4.07, p=0.014) | 1.41(0.63-3.17, p=0.4) |
| Consultation delay since the onset of symptoms (days) | >2 | 281 (63.6) | 33 (67.3) |  |  |
|  | ≤2 | 161 (36.4) | 16 (32.7) | 1.33(0.7-2.55, p=0.384) |  |
| SpO<sub>2</sub> level | <90% | 2 (0.4) | 4 (8.3) |  |  |
|  | [90,93%] | 38 (8.4) | 6 (12.5) | 17.17(2.29-128.54, p=0.006) | 3.38(0.21-53.25, p=0.386) |
|  | ≥94% | 410 (91.1) | 38 (79.2) | 26.33(4.24-163.68, p<0.001) | 4.50(0.35-58.11, p=0.249) |
| Severe malaria | No | 7 (1.5) | 2 (4.1) |  |  |
|  | Yes | 445 (98.5) | 47 (95.9) | 2.49(0.5-12.53, p=0.268) |  |
| Severe anaemia | No | 445 (98.5) | 31 (63.3) |  |  |
|  | Yes | 7 (1.5) | 18 (36.7) | 0.02(0.01-0.05, p<0.001) | 0.02(0.01-0.07, p<0.001) |
| **Severe acute malnutrition** | No | 429 (99.5) | 44 (97.8) |  |  |
|  | Yes | 2 (0.5) | 1 (2.2) | 0.14(0.01-1.74, p=0.128) | 0.43(0.01-14.77, p=0.64) |

### Day-14 mortality outcomes

At day-14, 2.2% (15/682) of severe pneumonia cases have died, 7.4% [9/122] among those hospitalised and 1.1% [6/560] among those not hospitalised. The proportion of death was significatively higher among those hospitalised compared to those not (p<0.001). Among the three children, whose accompanying persons have refused the HCW’s referral decision offered and returned to home, one child died and two were lost to follow-up.

## Discussion

We sought to describe the frequency, reasons and outcomes of the missed opportunities for hospitalisation of severe pneumonia identified with IMCI in PHC in four LMICs (Burkina Faso, Guinea, Mali, Niger) in West Africa. To the best of our knowledge, our study provides the first analysis of the missed opportunities of hospitalisation for IMCI-severe pneumonia diagnosed at PHC in West African settings where the prevalence of severe pneumonia among IMCI-severe cases aged 2-59 months was high, reaching 38% overall.

Our study highlights first the high rate of missed opportunities for hospitalisation of IMCI-severe pneumonia, reaching 82%, and mostly explained by the HCW decisions, rather than the family refusals. Thus, the fact is that this occurred mainly because HCW did not adhere to the IMCI severe pneumonia referral guideline recommending their systematic referral.

Previous studies in under-5 children have also reported a low adherence with IMCI guidelines referral regardless of the referral appropriateness [18–21]. This non-adherence to paediatric IMCI guidelines commonly reported in literature is explained by several reasons including the lack of training, lack of diagnostic tools, inadequate assessment, treatment, and monitoring, local clinical culture, infrastructure limitations, supply chain challenges, political context, family poverty, differences in patient-practitioner goals, disease trajectory, patient comorbidities, clarity and applicability of recommendations [22–24,24–29]. In our study, the high rate of missed opportunity of referral to hospital could be explained by several factors. Firstly, the lack of appropriation of the IMCI guidelines related to the lack of frequent MCI training of HCW in the PHC. Indeed, in the AIRE PHC, HCW reported that the IMCI trainings or refresh trainings were not enough regular regarding the frequent staff turn-over [11]. Despite the implementation of standardized guidelines, such as the IMCI guidelines or protocols for the management of specific pathologies, children remain not properly well-managed [30]. Secondly, the motivation of the HCW to follow the IMCI guidelines needs to be considered. Motivation is a central factor in determining adherence to health guidelines [31]. To improve the HCW motivation, it is important to conduct supervision and follow-up to reinforce adherence to guidelines over time. Thirdly, HCWs may have a good level of knowledge, but not applied it because they find it useless due to the lack of further health system support to refer and manage severe cases. In this context of high poverty, they do not want to expose households to high costs of care at district hospital level, as documented in the AIRE study [9]. These health expenses can be catastrophic for households. In these countries, the referral system could also be ineffective because of the lack of ambulance but also due to the geographical barriers, with long distances involved, and the security’ issues, particularly in Burkina Faso, Mali and Niger, as reported elsewhere [11]. Our data highlighted that when the HCW decided to refer a child, the vast majority of person accompanying the child adhered to it in our context, with only three family’ refusals of hospital referral who returned to home. Although we did not collect the reasons of these refusals, we assume they were related to the same poverty reasons as those of HCWs. Similarly, a study from rural Uganda reported lack of money, transport problems, and responsibilities at home as the main reasons declared by parents who refused hospital referral [32,33].

Second, when analysing the factors associated with missed opportunities of hospitalisation, our study also provides important findings regarding the HCW decision process, mainly guided by the clinical factors. First, HCW were more likely to refer the severe pneumonia complicated either by a severe anaemia, or a severe hypoxemia using PO. In the analysis restricted to Guinea cases, only severe anaemia reduced the odds of not being hospitalised without significant effect of SpO_2_ level. Thus, children who had SpO_2_<90% (except in Guinea) and with severe anaemia were undoubtedly perceived by HCW as being the sickest and consistently referred more frequently for care at hospital level. This is also indirectly confirmed by the significantly higher mortality at D-14 in the group hospitalised compared to those not, as marker of high severity. This result is in line with our findings reported elsewhere [12] and also other studies which reported that PO increased HCW confidence in their decision-making regarding child management [34,35]. However, we report that referral was significantly lower for severe pneumonia also complicated by severe malaria, which is worrying. This finding could be explained by the HCW self-perception of their experience to even manage severe malaria at the PHC level. In addition, the national malaria treatment guidelines differ on referral decision in cases of severe malaria. In Burkina Faso, severe malaria should be referred in cases of co-morbidity with severe anaemia, renal failure or coma but not explicitly severe pneumonia [13]. This gap in managing severe co-morbidity combining pneumonia and malaria needs to be specifically highlighted and mitigated in further guidelines and practices.

Finally, our findings also highlight the huge heterogeneity observed in the prevalence of severe pneumonia between the four countries, with Guinea bearing the most burden, as reported elsewhere. This result is explained by the difference in national IMCI protocols adapted from the WHO protocol. In Guinea, for the diagnosis of pneumonia, “Chest indrawing” is a sign of severity, whereas in Mali, Burkina Faso and Niger, this sign classifies children as moderate cases. In addition, Burkina Faso and Niger apply a total free care policy for all under-5 children years of age [36,37]. In these countries, assume that this policy has partly facilitated an earlier access to care for sick children leading to the lowest prevalence of severe pneumonia in these countries [38,39]. Some sick children also go directly to the hospital, without going through the PHC [40]. Mali and Guinea are countries that apply a policy of partial free healthcare for children under-5 [41–43] targeting four conditions (malaria, malnutrition, HIV and tuberculosis). In Guinea, such a policy may have contributed to increase the prevalence of severe cases including severe pneumonia by delaying access to primary care [9]. Geographical factors (relief, rivers), the absence of roads and long travel distances to reach PHC are additional barriers to access to health facilities in Guinea [11].

Our study has some limitations. The IMCI classification was done on the onsite HCW, that can lead to information bias. The IMCI guideline is a symptom-based guideline, that is operator-dependent, and can lead to inaccurate diagnostic in the absence of etiological diagnostic tools. In addition, the adaptation of the WHO standard guidelines in each country presents slightly differences, particularly in Guinea, and more specifically for the IMCI respiratory disease block. These differences in IMCI guidelines application are key factors to be considered when comparing the results between the different countries. The AIRE research project was also setup in some selected PHCs not representative of the national level. Our research teams were not present on-site at night, on weekends, or during public holidays, due to logistical and also security reasons, particularly in Mali and Burkina Faso. This selection bias may have conducted to underestimate the prevalence of severe pneumonia, assuming the fact that night and weekend attendance could concentrate the most severely ill. Therefore, our study population is probably not representative of all severe pneumonia cases in these West-African countries. Nevertheless, the strengths of our study are that data collection was standardised between countries, of good quality, with very few missing data overall, which made it possible to compare results between countries. We thus, provide original data on the rates of missed opportunities for hospitalisation of severe pneumonia and their main explanations in these West African settings. We also showed that the use of PO to diagnose severe hypoxemia has played a significant role in the HCW decision, adding evidence to its role. Finally, our data are useful for assessing the quality-of-care management of severe pneumonia at the PHC level in these West African settings, and for guiding revision of national policies.

## Conclusion

The proportion of severe pneumonia in under-5 children who were not referred to hospital was high in these primary care West African settings, and mainly related to the HCW’ decision. The hospitalisation of children with severe pneumonia was more frequent when complicated by severe anaemia or a diagnosed severe hypoxemia, highlighting a clinical consistency. However, those associated to severe malaria were less frequently referred, which needs to be further adressed. Strengthening the referral system, HCW training, supervision and follow-up to reinforce HCW adherence to guidelines over time is essential to support the appropriate referral of severe pneumonia and improve their survival and quality of care.

## Appendix

### ACKNOWLEDGMENTS

We thank all the children and their families who participated in the study, as well as the healthcare staff at the participating hospitals and the sites involved. We thank the field project staff and the AIRE Research Study Group*. We thank the Ministries of Health of the participating countries for their support.

**Acknowledgements: Children, families, UNITAID and The AIRE Research Study Group: Country investigators**: Ouagadougou, Burkina Faso: S. Yugbaré Ouédraogo (PI), V. M. Sanon Zombré (CoPI), Conakry, Guinea: M. Sama Cherif (CoPI), I. S. Diallo (CoPI), D. F. Kaba, (PI). Bamako, Mali: A. A. Diakité (PI), A. Sidibé, (CoPI). Niamey, Niger: H. Abarry Souleymane (CoPI), F. Tidjani Issagana Dikouma (PI). **Research coordinators & data centers: Inserm U1295, Toulouse 3 University, France:** H. Agbeci (Int Health Economist), L. Catala (Research associate), D. L. Dahourou (Research associate), S. Desmonde (Research associate), E. Gres (PhD Student), G. B. Hedible (Int research project manager), V. Leroy (research coordinator), L. Peters-Bokol (Int clinical research monitor), J. Tavarez (Research project assistant), Z. Zair (Statistician, Data scientist). **CEPED, IRD, Paris, France:** S. Louart (process manager), V. Ridde (process coordination). **Inserm U1137, Paris, France:** A. Cousien (Research associate). **Inserm U1219,** EMR271 IRD, **Bordeaux University, France**: R. Becquet (Research associate), V. Briand (Research associate), V. Journot (Research associate). **PACCI, CHU Treichville, Abidjan, Côte d’Ivoire**: S. Lenaud (Int data manager), C. N’Chot (Research associate), B. Seri (Supervisor IT), C. Yao (data manager supervisor). **Consortium NGOs partners: Alima-HQ (consortium lead), Dakar, Sénégal**: G. Anago (Int Monitoring Evaluation Accountability And Learning Officer), D. Badiane (Supply chain manager), M. Kinda (Director), D. Neboua (Medical officer), P. S. Dia (Supply chain manager), S. Shepherd (referent NGO), N. di Mauro (Operations support officer), G. Noël (Knowledge broker), K. Nyoka (Communication and advocacy officer), W. Taokreo (Finance manager), O. B. Coulidiati Lompo (Finance manager), M. Vignon (Project Manager). **Alima, Conakry, Guinea:** P. HAba (clinical supervisor), N. Diallo (clinical supervisor), M. Ngaradoum (Medical Team Leader), S. Léno (data collector), A. T. Sow (data collector), A. Baldé (data collector), A. Soumah (data collector), B. Baldé (data collector), F. Bah (data collector), K. C. Millimouno (data collector), M. Haba (data collector), M. Bah (data collector), M. Soumah (data collector), M. Guilavogui (data collector), M. N. Sylla (data collector), S. Diallo (data collector), S. F. Dounfangadouno (data collector), T. I. Bah (data collector), S. Sani (data collector), C. Gnongoue (Monitoring Evaluation Accountability And Learning Officer), S. Gaye (Monitoring Evaluation Accountability And Learning Officer), J. P. Y. Guilavogui (Clinical Research Assistant), A. O. Touré (Country health economist), J. S. Kolié (Country clinical research monitor), A. S. Savadogo (country project manager). **Alima, Bamako, Mali:** F. Sangala (Medical Team Leader), M. Traore (Clinical supervisor), T. Konare (Clinical supervisor), A. Coulibaly (Country health economist), A. Keita (data collector), D. Diarra (data collector), H. Traoré (data collector), I. Sangaré (data collector), I. Koné (data collector), M. Traoré (data collector), S. Diarra (data collector), V. Opoue (Monitoring Evaluation Accountability And Learning Officer), F. K. Keita (medical coordinator), M. Dougabka (Clinical research assistant then Monitoring Evaluation Accountability And Learning Officer), B. Dembélé (data collector then Clinical research assistant), M. S. Doumbia (country health economist), G. D. Kargougou (country clinical research monitor), S. Keita (country project manager). **Solthis-HQ, Paris**: S. Bouille (NGO referent), S. Calmettes (NGO referent), F. Lamontagne (NGO referent). **Solthis, Niamey:** K. H. Harouna (clinical supervisor), B. Moutari (clinical supervisor), I. Issaka (clinical supervisor), S. O. Assoumane (clinical supervisor), S. Dioiri (Medical Team Leader), M. Sidi (data collector), K. Sani Alio (Country supply chain officer), S. Amina (data collector), R. Agbokou (Clinical research assistant), M. G. Hamidou (Clinical Research Assistant), S. M. Sani (Country health economist), A. Mahamane, Aboubacar Abdou (data collector), B. Ousmane (data collector), I Kabirou (data collector), I. Mahaman (data collector), I Mamoudou (data collector), M. Baguido (data collector), R. Abdoul (data collector), A. Sahabi (data collector), F. Seini (data collector), Z. Hamani (data collector), L-Y B Meda (Country clinical research monitor), Mactar Niome (country project manager), X. Toviho (Monitoring Evaluation Accountability And Learning Officer), I. Sanouna (Monitoring Evaluation Accountability And Learning Officer), P. Kouam (program officer). **Terre des hommes-HQ, Lausanne:** S. Busière (NGO referent), F. Triclin (NGO referent). **Terre des hommes, BF:** A. Hema (country project manager), M. Bayala (IeDA IT), L. Tapsoba (Monitoring Evaluation Accountability And Learning Officer), J. B. Yaro (Clinical research assistant), S. Sougue (Clinical research assistant), R. Bakyono (Country health economist), A. G. Sawadogo (Country clinical research monitor), A. Soumah (data collector), Y. A. Lompo (data collector), B. Malgoubri (data collector), F. Douamba (data collector), G. Sore (data collector), L. Wangraoua (data collector), S. Yamponi (data collector), S. I. Bayala (data collector), S. Tiegna (data collector), S. Kam (data collector), S. Yoda (data collector), M. Karantao (data collector), D. F. Barry (Clinical supervisor), O. Sanou (clinical supervisor), N. Nacoulma (Medical Team Leader), N. Semde (clinical supervisor), I. Ouattara (Clinical supervisor), F. Wango (clinical supervisor), Z. Gneissien (clinical supervisor), H. Congo (clinical supervisor). **Terre des hommes, Mali:** Y. Diarra (clinical supervisor), B. Ouattara (clinical supervisor), A. Maiga (data collector), F. Diabate (data collector), O. Goita (data collector), S. Gana (data collector), S. Diallo (data collector), S. Sylla (data collector), D. Coulibaly (Tdh project manager), N. Sakho (NGO referent). **Country SHS team: Burkina Faso**: K. Kadio (consultant and research associate), J. Yougbaré (data collector), D. Zongo (data collector), S. Tougouma (data collector), A. Dicko (data collector), Z. Nanema (data collector), I. Balima (data collector), A. Ouedraogo (data collector), A. Ouattara (data collector), S. E. Coulibaly (data collector). **Guinea**: H. Baldé (consultant and research associate), L. Barry (data collector), E. Duparc Haba (data collector). **Mali**: A. Coulibaly (consultant and research associate), T. Sidibe (data collector), Y. Sangare (data collector), B. Traore (data collector), Y. Diarra (data collector). **Niger**: A. E. Dagobi (consultant and research associate), S. Salifou (data collector), B. Gana Moustapha Chétima (data collector), I. H. Abdou (data collector)

### FOOTNOTES

#### Handling editor

Contributors: VL and VR conceptualised the AIRE research protocol. The AIRE Research Study Group conducted training, data collection and management. ZZ with contributions from DD, DN, HGB and VL conceptualised and realised this data analysis. DD wrote the first draft of this article. All authors were involved in data interpretation and review of the final manuscript. VL is the guarantor to submit the manuscript.

#### Funding

The AIRE project is funded by UNITAID, with in-kind support from Inserm and IRD. UNITAID was not involved in the design of the study, the collection, analysis and interpretation of the data, nor in the writing of the manuscript.

#### Competing interest

All authors have declared no conflict of interest.

#### Ethics approval and consent to participate

Ethics approval and consent to participate The AIRE research protocol, the information notice (translated in vernacular languages), the written consent form and any other relevant document have been submitted to each national ethics committee, to the Inserm Institutional Evaluation Ethics Committee (IEEC) and to the WHO Ethics Review Committee (WHO ERC). All the aforementioned ethical committees reviewed and approved the protocol and other key documents (Comité d’Ethique pour la Recherche en Santé (CERS), Burkina Faso n°2020–4 070; Comité National d’Ethique pour la Recherche en Santé (CNERS), Guinea n°169/CNERS/21; Comité National d’Éthique pour la Santé et les Sciences de la vie (CNESS), Mali n°127/MSDS CNESS; Comité National d’Ethique pour la Recherche en Santé (CNERS) Niger n°67/2020/CNERS; Inserm IEEC n°20–720; WHO ERC n° ERC.0003364). This study has been retrospectively registered by the Pan African Clinical Trials Registry on June 15th 2022 under the following Trial registration number: PACTR202206525204526.

#### Data Availability Statement

The datasets generated and analysed during the current study are not publicly available. Access to processed deidentified participant data will be made available to any third Party after the publication of the main AIRE results stated in the Pan African Clinical Trial Registry Study statement (PACTR202206525204526, registered on 06/15/2022), upon a motivated request (concept sheet), and after the written consent of the AIRE research coordinator (Valeriane Leroy, Inserm U1295 Toulouse, France) obtained after the approval of the AIRE publication committee, if still active.

## Notes

### Competing Interest Statement

The authors have declared no competing interest.

### Clinical Trial

PACTR202206525204526

### Funding Statement

The AIRE project was funded by UNITAID, with in-kind support from Inserm and IRD.

### Author Declarations

– The WHO Ethics Review Committee (WHO-ERC) gave ethical approval for this work (ERC.0003364) – The Inserm Institutional Evaluation Ethics Committee (IEEC) gave ethical approval for this work (Inserm IEEC 20-720). – The "Comite d Ethique pour la Recherche en Sante (CERS), of Burkina Faso gave ethical approval for this work (2020-4-070). – The "Comite National d Ethique pour la Recherche en Sante (CNERS)", of Guinea gave ethical approval for this work (169/CNERS/21). – The "Comite National d Ethique pour la Sante et les Sciences de la vie (CNESS)", of Mali gave ethical approval for this work (127/MSDS-CNESS). – The "Comite National d Ethique pour la Recherche en Sante (CNERS)", of Niger gave ethical approval for this work (67/2020/CNERS).

